## Supplementary Text file for "Human leishmaniasis vaccines: use cases, target population and potential global demand"

### Supplementary Text to the Article “Human Leishmaniasis Vaccines: use cases, target population and potential global demand”

#### DEFINTION OF EPIDEMIOLOGICAL ESTIMATES

##### A. Estimate of population at high risk of VL in 2018 – used in forecasting demand for prophylactic VL vaccine

Step 1: identification of alternative sources for Global population at risk:

- WHO country profiles[i] integrated with Pigott et al. 2014[ii] for year 2014 – the estimate includes 14 countries identified by WHO as high burden countries and 11 others with incidence above 1/100k according to the GBD 2017[iii]. This selection aims at reflecting countries where VL could be considered a sufficiently high public health burden to justify the investment into the adoption of a vaccine. Data for 13 countries reflect WHO estimates from Leishmaniasis relevant country profiles; the other 12 countries are sourced from Pigott estimates.

| | | | | | | | | | Estimated Population at Risk of VL | | | 2019 Visceral leishmaniasis national incidence per 100,000, all ages, (GBD 2019) | | |
|---|---|---|---|---|---|---|---|---|---|---|---|---|---|---|
| Nr. | Country | WHO HIGH RISK | REGION | WHO REGION | WB Income Level (2020) | GAVI SUPPORT | DISEASE | VL TRANSMISSION | WHO Estimates | Other Estimates | Pigott 2014 | val | upper | lower |
| 1 | **South Sudan** | X | AFRICA | EMRO | LIC | X | VL | ANTHROP. | 2'034'944 | | 3'749'817 | 46.57 | 68.28 | 30.82 |
| 2 | **Sudan** | X | AFRICA | AFRO | LIC | X | CL & VL | ANTHROP. | 8'696'636 | | 16'259'580 | 9.63 | 15.05 | 5.59 |
| 3 | **Somalia** | X | AFRICA | EMRO | LIC | X | VL | ANTHROP. | 2'377'787 | | 2'363'005 | 6.88 | 10.13 | 4.55 |
| 4 | **Ethiopia** | X | AFRICA | AFRO | LIC | X | VL | | 3'168'835 | | 36'732'612 | 1.95 | 2.89 | 1.28 |
| 5 | **Kenya** | X | AFRICA | AFRO | LMIC | X | VL | | 3'268'626 | | 14'151'164 | 2.16 | 2.93 | 1.56 |
| 6 | **Brazil** | X | AMERICA | PAHO | UMIC | | CL & VL | | 80'235'408 | | 103'739'410 | 2.30 | 3.31 | 1.57 |
| 7 | **Georgia** | X | EUROPE | EURO | UMIC | X | VL | | 2'593'595 | | 1'799'810 | 2.04 | 3.13 | 1.17 |
| 8 | **Nepal** | X | ASIA | SEARO | LMIC | X | VL | ANTHROP. | 28'624'296 | | 13'864'455 | 0.92 | 1.41 | 0.57 |
| 9 | **India** | X | ASIA | SEARO | LMIC | X | CL & VL | ANTHROP. | 130'000'000 | | 495'733'890 | 0.59 | 0.92 | 0.34 |
| 10 | **Bangladesh** | X | ASIA | SEARO | LMIC | X | VL | ANTHROP. | 30'000'000 | | 72'436'664 | 0.22 | 0.34 | 0.13 |
| 11 | **Spain** | X | EUROPE | EURO | HIC | | VL | | 37'154'139 | | 31'302'114 | 0.12 | 0.20 | 0.06 |
| 12 | **China** | X | ASIA | WPRO | UMIC | | VL | PARTIALLY | 229'718'479 | | 205'894'780 | 0.06 | 0.08 | 0.05 |
| 13 | **Paraguay** | X | AMERICA | PAHO | UMIC | | CL & VL | | 3'060'725 | | 2'764'511 | 0.82 | 1.17 | 0.56 |
| 14 | **Uganda** | X | AFRICA | AFRO | LIC | X | VL | | | | 4'603'301 | 0.22 | 0.36 | 0.12 |
| 15 | **Central African Republic** | | AFRICA | AFRO | LIC | X | VL | | | | 1'069'348 | 7.61 | 26.60 | 1.08 |
| 16 | **DR Congo** | | AFRICA | AFRO | LIC | X | VL | | | | 12'956'326 | 3.26 | 11.74 | 0.50 |
| 17 | **Malawi** | | AFRICA | AFRO | LIC | X | VL | | | | 4'266'093 | NA | NA | NA |
| 18 | **Niger** | | AFRICA | AFRO | LIC | X | VL | | | | 2'398'483 | 2.24 | 7.84 | 0.32 |
| 19 | **Tanzania** | | AFRICA | AFRO | LMIC | X | VL | | | | 1'575'703 | NA | NA | NA |
| 20 | **Eritrea** | | AFRICA | AFRO | LIC | X | VL | | | | 2'665'204 | 2.36 | 5.29 | 0.83 |
| 21 | **Chad** | | AFRICA | AFRO | LIC | X | VL | | | | 2'679'550 | 1.11 | 3.51 | 0.18 |
| 22 | **Zambia** | | AFRICA | AFRO | LMIC | X | VL | | | | 4'129'548 | 1.22 | 4.43 | 0.18 |
| 23 | **Angola** | | AFRICA | AFRO | LMIC | X | VL | | | | 1'276'322 | 1.55 | 4.99 | 0.26 |
| 24 | **Kyrgyzstan** | | ASIA | EURO | LMIC | X | VL | | | | 3'003'517 | 1.12 | 3.87 | 0.19 |
| 25 | **Djibouti** | | AFRICA | EMRO | LMIC | X | VL | | | | 652'505 | 4.31 | 6.96 | 2.55 |

- WHO TRS 2010[iv] for 2010 – the estimate includes global population at risk for all leishmaniases. The totality of this undifferentiated population at risk has been attributed to VL, consistently with the detailed Pigott estimates where the difference between VL and CL at risk populations was minimal.
- DnDI newsletter 2009[v] - global estimate for Visceral Leishmaniasis for 2009

Step 2: definition of 2018 estimates across the different sources

In order to define comparable estimates, projections of older data point have been created applying the population growth rate for the period 2010-2015 as reported by UN/DESA. The growth rate is a weighted growth rate based on number of VL cases for 2019 as per the Global Burden of Diseases 2019 in the regions where VL is endemic (Asia, Africa and Mediterranean area, Americas). **The value used is 1.80% yearly growth.** No significant epidemiological change has been assumed.

Step 3: definition of 2018 estimate ranges

Based on the estimates resulting from step 2, the range for the global population at risk of VL has been defined as in the below table:

| Scenario | Source | 2018 Value |
|---|---|---|
| Upper limit | WHO country estimates and Pigott 2014 | **646 million** |

| | | |
|---|---|---|
| Base Case | WHO TRS 2010 | **404 million** |
| Lower Limit | DnDi 2009 | **235 million** |

Step 4: definition of future projections (period 2019-2040)
Starting from the 2018 data point, estimates of the population at risk for the period 2019-2040 have been created by applying a **1.44%** weighted population yearly growth rate for the period 2020-2040 as estimated by UN/DESA. The growth rate is a weighted growth rate based on number of cases in 2019 as per the Global Burden of Diseases 2019 in the regions where VL is endemic (Asia, Africa and Mediterranean area, Americas). No significant epidemiological change has been assumed for the future.

Step 5: definition of the impact of the introduction of a VL prophylactic vaccine on the population at risk of VL
It is estimated that the introduction of a VL prophylactic vaccine will only impact the size of the population at risk in areas with anthroponotic-only transmission in Asia (India, Bangladesh, Nepal and 19% of the total at risk population in China[vi]) and in Africa[vii] (Sudan and Somalia). In those areas the decline of the population is assumed to happen linearly over 4 years starting from 2035 (5 years after the start of the deployment of the vaccine when the entire population below 20 years is reached by the vaccine) under the assumption that those countries will progressively stop treatments/ immunization once all districts have reached elimination (e.g., having achieved reduction of cases below a defined incidence threshold). The total population at risk of the five countries above accounts for 245.5million or approximately 40% of the total population at risk as per country estimates from WHO.

#### B. Estimate of global VL incidence – year 2019 - used in forecasting demand for a preventative PKDL vaccine

Step 1: definition of 2019 estimated regional ranges for the PKDL endemic regions
Estimates are based on the 2019 ranges from the GBD 2019 for the 9 PKDL endemic countries:

| 9 countries |
|---|
| Africa: South Sudan, Sudan, Somalia, Eritrea, Ethiopia, Djibouti,<br>Asia: Nepal, India, Bangladesh |

The regional ranges result as follow:

| Region | Estimate | Source | 2019 Value |
|---|---|---|---|
| Asia | High | GBD 2019 | 13801 |
| | Medium | | 8771 |
| | Low | | 5104 |
| Africa | High | GBD 2019 | 18091 |
| | Medium | | 11959 |
| | Low | | 7351 |
| TOTAL 2019 | **High** | | **31892** |
| | **Medium** | | **20730** |
| | **Low** | | **12635** |

Step 2: definition of future projections.
The 2019 estimates have been projected through 2040 by taking into account the prevalent population growth rate for the period 2010-2019 from GBD 2019 (differentiated per region and per high/medium/low scenario).

| Region | CAGR 2010-2019 (Mid-point only) |
|---|---|
| Asia | -19.41% |
| Africa | -9.72% |

For the high and medium scenarios, a “wave” trend is used whereby 5 years of decline are followed by 5 years of increase at the same growth rate that leads the prevalence to reach the same point after 10 years. 2019 is assumed to be in the middle of the declining wave. For the low scenario, a continued decline is instead assumed.

Step 3: definition of the impact of introduction of a VL prophylactic vaccine on VL incidence
In order to assess the impact of the introduction of a VL prophylactic vaccine, the results of the impact modelling performed by Erasmus Medical School for India (fully anthroponotic transmission) has been used. In such model, under the assumption that asymptomatic population also contributes to transmission, the reduction in number of cases following vaccination with a 50% efficacious vaccine (population is 50% less likely to get infected) and capable of covering 100% of the population is the following:

| | Year 1 | Year 2 | Year 3 |
|---|---|---|---|
| Reduction of cases | -30% | -50% | -62% |
| As % of vaccine efficacy | -60% | -100% | -124% |

On those bases, and taking into account (a) the year of introduction of the vaccine, and (b) the proportion of total population reached as result of the introduction sequence based on an EPI introduction, the following curve has been applied to estimate the reduction in the VL cases in each of the 3 scenarios:

| Year 1 | Year 2 | Year 3 | Year 4 | Year 5 | Year 6 | Year 7 | Year 8 | Year 9 |
|---|---|---|---|---|---|---|---|---|
| 2.8% | 16.5% | 31.9% | 49.6% | 65.6% | 77.8% | 85.5% | 91.9% | 95.0% |

The reduction in number of cases is applied with one-year delay compared to the year of introduction of the vaccine.

#### C. Estimate of global PKDL incidence – used in forecasting demand for PKDL therapeutic vaccine

Step 1: definition of the appropriate VL sequelae rate
Starting from the estimate of the VL cases range and projections as indicated above, the following sequelae rate are applied, calculated as average of the regional upper and lower limits

| Region | Source | High Sequelae rate | Low Sequelae rate | Average |
|---|---|---|---|---|
| Africa | P.Kaye 2019[viii] | 20% | 25% | 22.5% |
| Asia | Zijstra 2016[ix] | 10% | 20% | 15% |

Step 2: definition of the PKDL incidence range
As result the following ranges of PKDL incidence are estimated:

| Region | Estimate | 2019 Value |
|---|---|---|
| Africa | High | 4070 |

| | | |
|---|---|---|
| | Medium | 2691 |
| | Low | 1694 |
| asia | High | 2070 |
| | Medium | 1316 |
| | Low | 766 |
| TOTAL 2019 | **High** | **6141** |
| | **Medium** | **4006** |
| | **Low** | **2460** |

Step 3: <u>definition of the impact of introduction of a preventative PKDL vaccine on PKDL incidence</u>
Based on an assumption of years roll out of the vaccine to spread over 2 years and on the reduction of cases illustrated for VL, the following curve has been used to estimate the reduction in the number of cases in each of the 3 scenarios:

| Year | Year 1 | Year 2 |
|---|---|---|
| Reduction | 50% | 90% |

The reduction in number of cases is applied with one-year delay compared to the year of introduction of the vaccine (2027).

Step 4: <u>Estimate of global PKDL prevalence</u>
Starting from the estimate of PKDL incidence range as indicated above, an average duration of PKDL lesions of 3.5 years (based on Mondal 2018[x]) is applied to calculate the total number of active cases in each single year (e.g., the 2021 value will be equal to the sum of the number of new cases in 2018 (half year), 2019, 2020 and 2021.

| Region | Estimate | 2021 Value |
|---|---|---|
| Africa | High | 12911 |
| | Medium | 8658 |
| | Low | 5466 |
| Asia | High | 6278 |
| | Medium | 3888 |
| | Low | 2201 |
| TOTAL 2019 | **High** | **19189** |
| | **Medium** | **12547** |
| | **Low** | **7668** |

#### D. Estimate of population at high risk of CL – used for the forecasting of demand for a CL prophylactic vaccine

<u>Step 1: comparison of different sources:</u>

- WHO country profiles integrated with Pigott et al. 2014 for year 2014 – The estimates include 11 countries identified by WHO having high burden and 22 others with incidence above 5/100k according to the GBD 2019. India (2 states) have also been added taking the total to 34. This selection aims at reflecting countries where CL could be considered a sufficiently high public health burden to justify the investment into the adoption of a vaccine. Data for 10 countries and the total for the 14 countries in the PAHO region reflect WHO detailed estimates from Leishmaniasis relevant country/regional profiles. Other 9 countries are sourced from Pigott estimates, while for India the estimate corresponds to the population of Rajasthan and Kerala where the diseases is endemic.

| | | | | | | | | Estimated Population at Risk of CL | | | 2019 Cutaneous and mucocutaneous leishmaniasis - national incidence per 100,000, all ages, (GBD 2019) | | |
|---|---|---|---|---|---|---|---|---|---|---|---|---|---|
| Nr. | Country | WHO High Risk | REGION | WHO REGION | WB Income Level (2020) | GAVI SUPPORT | DISEASE | WHO Estimates | Other Estimates | Pigott 2014 | val | upper | lower |
| 1 | **Syria** | x | ASIA | EMRO | LIC | X | CL | 18'502'000 | | 20'784'102 | 728.63 | 1'409.58 | 258.37 |
| 2 | **Afghanistan** | x | ASIA | EMRO | LIC | X | CL | 10'340'735 | | 15'616'552 | 413.90 | 801.91 | 145.24 |
| 3 | **Tunisia** | x | AFRICA | EMRO | LMIC | | CL | 6'018'090 | | 9'711'311 | 152.72 | 304.52 | 51.41 |
| 4 | **Suriname** | | AMERICA | PAHO | UMIC | | CL | | | 520'165 | 149.60 | 183.56 | 116.94 |
| 5 | **Costa Rica** | | AMERICA | PAHO | UMIC | | CL | | | 4'243'499 | 126.91 | 259.81 | 43.76 |
| 6 | **Nicaragua** | | AMERICA | PAHO | LMIC | X | CL | | | 4'003'127 | 119.08 | 212.65 | 53.75 |
| 7 | **Panama** | | AMERICA | PAHO | HIC | | CL | | | 3'308'074 | 103.38 | 353.92 | 4.03 |
| 8 | **Libya** | | AFRICA | EMRO | UMIC | | CL | | | 5'716'142 | 98.19 | 218.08 | 25.87 |
| 9 | **Iraq** | | ASIA | EMRO | UMIC | | CL | | | 29'738'302 | 84.96 | 184.35 | 26.00 |
| 10 | **Yemen** | | ASIA | EMRO | LIC | X | CL | | | 20'909'202 | 80.87 | 162.36 | 27.19 |
| 11 | **Honduras** | | AMERICA | PAHO | LMIC | X | CL | | | 5'488'859 | 77.81 | 116.28 | 43.24 |
| 12 | **Algeria** | x | AFRICA | EMRO | LMIC | | CL | 10'005'224 | | 30'969'660 | 66.29 | 130.30 | 23.41 |
| 13 | **Peru** | x | AMERICA | PAHO | UMIC | | CL | | | 20'095'782 | 65.67 | 106.54 | 31.80 |
| 14 | **Bolivia** | | AMERICA | PAHO | LMIC | X | CL | | | 5'727'962 | 61.50 | 86.39 | 41.19 |
| 15 | **Colombia** | x | AMERICA | PAHO | UMIC | | CL | | | 44'869'432 | 55.09 | 197.48 | 1.50 |
| 16 | **Brazil** | x | AMERICA | PAHO | UMIC | | CL & VL | 240'000'000 | | 148'786'750 | 36.04 | 51.57 | 23.46 |
| 17 | **Morocco** | x | AFRICA | EMRO | LMIC | | CL | 6'130'393 | | 30'282'374 | 27.15 | 56.05 | 9.58 |
| 18 | **Ecuador** | | AMERICA | PAHO | UMIC | | CL | | | 12'469'102 | 26.75 | 54.11 | 8.99 |
| 19 | **West Bank** | | ASIA | EMRO | LMIC | | CL | | | 2'842'185 | 26.75 | 54.11 | 8.99 |
| | **Gaza Strip** | | ASIA | EMRO | LMIC | | CL | | | 1'515'670 | 26.40 | 60.81 | 6.71 |
| 20 | **Saudi Arabia** | x | ASIA | EMRO | HIC | | CL | 3'590'391 | | 21'682'114 | 24.49 | 45.28 | 10.35 |
| 21 | **Venezuela** | | AMERICA | PAHO | UMIC | | CL | | | 26'249'248 | 23.96 | 38.01 | 12.50 |
| 22 | **Burkina Faso** | | AFRICA | AFRO | LIC | X | CL | | | 5'379'299 | 23.21 | 42.76 | 9.40 |
| 23 | **Guyana** | | AMERICA | PAHO | UMIC | X | CL | | | 651'271 | 16.93 | 76.68 | 0.04 |
| 24 | **Turkmenistan** | | ASIA | EURO | UMIC | | CL | | | 3'957'351 | 14.82 | 38.10 | 2.19 |
| 25 | **Sri Lanka** | | ASIA | SEARO | LMIC | X | CL | | | 7'250'986 | 12.25 | 36.61 | 0.95 |
| 26 | **Guatemala** | | AMERICA | PAHO | UMIC | | CL | | | 8'991'769 | 10.23 | 41.95 | 0.13 |
| 27 | **Jordan** | | ASIA | EMRO | UMIC | | CL | | | 6'358'596 | 8.08 | 16.96 | 2.36 |
| 28 | **Turkey** | x | ASIA | EURO | UMIC | | CL | 41'658'616 | | 20'876'026 | 7.46 | 16.74 | 2.18 |
| 29 | **Sudan** | | AFRICA | AFRO | LIC | X | CL & VL | 37'419'625 | | 21'327'576 | 7.64 | 16.78 | 2.06 |
| 30 | **Uzbekistan** | | ASIA | EURO | LMIC | X | CL | 15'491'744 | | 12'793'034 | 7.66 | 17.08 | 1.63 |
| 31 | **Paraguay** | | AMERICA | PAHO | UMIC | | CL & VL | | | 5'209'033 | 7.56 | 15.26 | 2.52 |
| 32 | **Pakistan** | | ASIA | EMRO | LMIC | X | CL | 86'430'000 | | 156'427'700 | 6.80 | 10.21 | 3.99 |
| 33 | **Iran** | x | ASIA | EMRO | UMIC | | CL | | | 60'143'656 | 3.64 | 7.21 | 1.47 |
| 34 | **India (2 states)** | | ASIA | SEARO | LMIC | X | CL & VL | | 104'000'000 | 319'099'420 | 0.03 | 0.04 | 0.02 |

- DnDI Disease Profile 2018[xi] for all cases of leishmaniasis (assumed 100% for CL)
- WHO TRS 2010[xii] for 2010 for all leishmaniasis (assumed 100% for VL)

Step 2: definition of 2018 estimates across the different sources

In order to define comparable estimates, projections of older data point have been created applying the population growth rate for the period 2010-2015 as reported by UN/DESA. The growth rate is a weighted growth rate based on number of 2019 CL cases as per the Global Burden of Diseases 2019 in the regions where CL is endemic (Asia, Africa and Mediterranean area, Americas).). **The value used is 1.27% yearly growth.** No significant epidemiological change has been assumed.

Step 3: definition of 2018 estimate ranges

For the global population at risk the following data are selected:

| Scenario | Source | 2018 Value |
|---|---|---|
| Upper limit | DnDI 2018 | **1 billion** |
| Base Case | WHO country estimates and Pigott 2014 | **773 million** |
| Lower Limit | WHO TRS 2010 | **399 million** |

Step 4: definition of future projections

Starting from the 2018 data point, estimates of the population at risk for the period 2019-2040 have been created by applying a **1.27%** weighted population yearly growth rate for the period 2020-2040 as estimated by UN/DESA. The growth rate is a weighted growth rate based on number of CL cases in 2019 as per the Global Burden of Diseases 2019 in the regions where CL is endemic (Asia, Africa and Mediterranean area, Americas). No significant epidemiological change has been assumed for the future.

Step 5: definition of the impact of introduction of a vaccine on the population at risk of CL

It is estimated that the vaccine will not have any impact in the reduction of the population at risk of CL the disease being a zoonosis (with the exception of L. tropica)

#### DEFINTION OF VACCINE AND FINANCIAL PARAMETERS

##### E. Definition of Vaccine parameters

Age of administration of the first dose of the vaccine is defined based on the TPP (with no specific age for PKDL) determining the time of the first and eventually subsequent administration.
Efficacy of the vaccine (in term of reduction of number of cases) is defined based on the TPP
Duration of protection of the vaccine is defined based on the TPP informing the number of series required for the vaccine
Number of doses per series is defined based on the TPP
Number of required series is calculated based on the duration of protection and limited to a maximum of 3 series. No vaccine is currently administered with more than 4 series (DTP 3 doses in the first year of life and 3 boosters between 4 and 15 years of age).

| Indication | Age for first dose | Efficacy | Duration of Protection | Nr. Doses per Series | Series |
|---|---|---|---|---|---|
| VL prophylactic | 1 year | 70-95% | 5 years | 2 | 3 (at year 1, 6 and 11) |
| CL prophylactic | 1 year | 70-90% | 5 years | 2 | 3 (at year 1, 6 and 11) |
| VL/CL pro catch-up | | 70-90% | | 2 | 1 |
| PKDL therapeutic | NA | 30-90% | Lifelong | 1 | 1 |
| PKDL preventive | NA | 30-90% | Lifelong | 1 | 1 |

Year of licensure and of prequalification (differentiated for VL, CL and PKDL). Under the assumption that the vaccine will be manufactured and commercialized by an experienced manufacturer familiar with the prequalification processes, a delay of 12 months has been assumed for the time of achievement of the WHO PQ from the time of the first registration.

| Indication | Year of first registration | Year of WHO prequalification |
|---|---|---|
| VL prophylactic | 2029 | 2030 |
| CL prophylactic | 2029 | 2030 |
| PKDL therapeutic | 2027 | NA |
| PKDL preventive | 2027 | NA |

##### F. Definition of Implementation parameters

Coverage proxies are defined for the different modes of delivery:

- Coverage proxy for routine delivery in the first year of life is defined based on WHO & UNICEF Coverage Estimates WUENIC[xiii] estimates of DTP1 and 3 and MCV1 for the African region
- Coverage proxy for routine delivery via school administration (5 to 14 years of life) is based on HPV coverage estimates (as per Bruni 2016[xiv])
- Coverage proxy for routine adult and out-of-school delivery is based on anecdotal evidence from WHO
- Coverage proxy for campaign delivery is based on anecdotal evidence from Gavi and WHO

Primary and Secondary school enrolment rates are based on UNICEF estimates[xv] for the African region and used to calculate the coverage of the population in the school period according to this formula:

AVERAGE ((School coverage * Primary Enrollment rate + Out-of-school coverage * (1 – Primary Enrollment rate), (School coverage * Secondary Enrollment rate + Out-of-school coverage * (1 – Secondary Enrollment rate))

Definition of Leishmaniasis vaccine target coverage
On those bases, relevant target coverage data points – e.g. the coverage that will be reached by the vaccine once fully introduced in the national immunization program of a country - are defined depending on the administration age/es resulting from the assumptions in term of age of first administration, duration of protection and number of series.

| Indication | Delivery | Proxy | Coverage |
|---|---|---|---|
| VL & CL Prophylactic | Routine at 9 months-2$^{nd}$ year of life | MCV1 | 73% |
| | Routine at 6 years – 11 years | HPV | 68% |
| | Routine for adults | Anecdotal | 45% |
| | Campaigns delivery (including catch-up) | SIA analyses | 90% |
| PKDL Therapeutic & Preventive | Following delivery of VL or PKDL treatment | 100% in Indian Subcontinent 85% in Africa | 93% |

Definition of the standard sequence of country vaccine routine introductions (uptake curve) – looking at the status of the countries most likely to introduce the vaccine, 20 of the 26 countries (77%) likely target for a VL vaccine introduction and 14 of the 36 countries (39%) likely target for a CL vaccine are or were eligible for Gavi support. The sequence of introduction of PCV (only vaccine rolled-out in the largest majority of countries with no relevant supply of funding constraints) in the Gavi countries is selected as the one providing the most accurate proxy for the sequence (number of country introductions per year) of introductions into routine immunization in the targeted countries. The curve indicates the growing share of the targeted population reachable by the vaccine as result of country decisions to introduce the vaccine in their immunization schedule.

Wastage rate: based on WHO standard guidance for a low-multidose (5 or 10 doses) presentation
Buffer stocks: as per Gavi operational forecast: every year 25% of change in total demand
Price per dose: 2.5 USD (or 5 doses per course) as per the TPP
Cost of Goods (COGs): 1 USD as per the TPP

#### G. Definition of financial parameters

Sales, general and administrative expenses (SG&A): 8.6% equal to half of reported expenses by a large corporation (Sanofi 2017)
Depreciation rate for the plant: 15 years based on GAAP rules
Taxation rate: not knowing the location of the manufacturer and the legal structure for commercialization the average worldwide corporate tax rate of 23% is used as per Tax Foundation data set
Weighted Average Cost of Capital: 8.7% for pharmaceutical and biotech companies as per NYU Stern business school[xvi]

**DOSES CALCULATION**

#### H. Definition of target population for different indications and delivery strategy

Definition of the target population for catch-up campaigns for the prophylaxis against VL and CL
It is assumed that each country will perform a catch-up campaign at start of the program to reach a portion of the population at risk that will not be otherwise reached with the start of routine

immunization (because of older age). On the base of the age distribution of the cases (as per GBD 2017) the following target populations are assumed for catch-up:

| Indication | Age distribution | Target age for Catch-Up |
|---|---|---|
| Prophylaxis of VL | Based on VL cases:<br>30% - 0 to 4 years<br>35% - 5 to 14 years<br>30% - 15 to 49 years<br>5% - 50 years and above | **5 to 14 years**<br>Catch-up in school to be followed by regular start in EPI at 1 year of age |
| Prophylaxis of CL | Based on CL cases<br>2% - 0 to 4 years<br>15% - 5 to 14 years<br>59% - 15 to 49 years<br>24% - 50 years and above | **5 to 29 years**<br>SIA to be followed by start in EPI at 1 year of age |

<u>Definition of the target population for routine immunization</u>
For each year from 2018 through 2040, the population targeted by the vaccine under the various indications is defined as follow:

- **VL**: the total population at risk of VL has been split in age groups based on age distribution as per UN/DESA to be able to determine the size of the age groups that will be targeted by the selected routine vaccination strategy and the appropriate coverage level/s (see above the implementation parameter section). Based on the schedules (number of series) indicated in the TPP it is assumed that one first cohort (at 1 year of age) will be targeted in the 0 to 4 years of age group, if needed two cohorts (at 6 at 11 years of age) will be targeted in the 5 to 14 years of age group and if needed two cohorts (at 16 and 21 years of age) will be targeted in the older age. Cohort populations are assumed to be equally distributed within the age range.
- **CL**: the total population at risk of CL has been split in age groups based on age distribution as per UN/DESA to be able to determine the size of the age groups that will be targeted by the selected routine vaccination strategy and the appropriate coverage level/s (see above the implementation parameter section). **Based on the targeted countries population at risk a reduction factor of 28.7% is applied to reflect the populations that will be already reached for VL vaccination in India, Sudan, Brazil & Paraguay**. Based on the schedule (number of series) indicated in the TPP it is assumed that one first cohort (at 1 year of age) will be targeted in the 0 to 4 years of age group, if needed two cohorts (at 6 at 11 years of age) will be targeted in the 5 to 14 years of age group and if needed two cohorts (at 16 and 21 years of age) will be targeted in the older age. Cohort populations are assumed to be equally distributed within the age range.

| | Population distribution (UN/DESA) |
|---|---|
| PSAC (0-4 years) | 9% |
| SAC (5-14 years) | 18% |
| Adolescents & Young adults (15-29 years) | 25% |
| Adults and Elderly (above 29 years) | 48% |

<u>Definition of the target population in the treatment and prevention of PKDL:</u>

- **PKDL Therapeutic**: under the assumption that the total PKDL affected population will be targeted, the calculated global number (see above) is considered
- **PKDL Preventive**: under the assumption that the total VL cases will be targeted, the calculated global number (see above) is considered

#### I. Calculation of doses required

Without considering the time of introduction of the vaccine in the different indication or the programmatic requirements, for each year from 2018 through 2040, the following calculations are performed

- Calculation of **"coverable" population**:
    - VL and CL catch up: for the age groups targeted (as defined above) the SIA coverage is applied to define the maximum number of people that will likely be reachable by the intervention as per the formula:
      "target population x coverage"
    - VL and CL routine: for each cohort the appropriate coverage (as defined above) is applied to the target population to define the number of people that will likely be reachable by the intervention as per the formula:
      "target population x coverage"

  - For PKDL: a delivery post VL/PKDL treatment is foreseen, hence no age specific coverage will be applied but the general leishmaniasis treatment coverage as per the formula:
    "target population x coverage"

- Calculation of **100% Administrable doses:**
    - Based on the number of series required in each delivery strategy/indication and on doses per series (as defined above) and on the coverable population (as defined above) the number of doses required to vaccinate the population that can be reached by the immunization system is calculated as per the formula:
      "coverable population x number of series x doses per series"

#### J. Calculation of doses employed

For each indication, considering the time of introduction of the vaccine, the number of doses required is calculated as follow

- Definition of the country uptake:
    - For the catch-up (VL & CL only) the activity is one-off therefore the country uptake curve (as defined above) provides, via the calculation of the yearly increase (vs. the cumulative number), the portion of the total population that is caught in a specific year.
    - For the routine program (VL & CL only) the country uptake curve (as defined above) is applied for each of the indications based on the year of registration (as defined above) defining the proportion of the 100% administrable doses that will be reached. The uptake curve is applied unmodified for the PSAC that are reached every year since the introduction of the vaccine and with a delay for the two administrations foreseen for SAC to discount the fact that catch-up campaigns initially cover those populations. The delay accounts for the fact that the first booster will be administered from year 3 and the second booster from year 5 and results in the following modified uptake curve for PSAC:

| | | | | | | | | | | | |
|---|---|---|---|---|---|---|---|---|---|---|---|
| 0% | 0% | 2% | 13% | 22% | 44% | 56% | 74% | 83% | 94% | 96% | 100% |

  - For the ongoing prevention and treatment of PKDL, the country uptake curve (as defined above) is applied for each of the indications based on the year of registration (as defined above) defining the proportion of the 100% administrable doses that will be reached.

- Calculation of the doses administered: based on the selected uptake curve the total number of doses administered is calculated for each year according to the following formula: "uptake x target population"

- Calculation of the wastage: for each year the standard % wastage rate (as defined above) is applied to the administered doses according to the following formula: "administered doses x (1 + wastage rate)"

- Calculation of the buffer stock: for each year the required buffer stock is calculated according to the formula: "administered doses year$^{x}$ – administered doses year$^{(x-1)}$ x buffer stock rate"

- Calculation of the total number of doses required: for each year, the total number of doses whose production will be required to fulfill the emerging demand is calculated according to the following formula: "administered doses + wastage + buffer stock"

**DISCOUNTED CASH FLOW ANALYSIS**

The Discounted Cash Flow (DCF) is the standard methodology used to compare positive and negative flows of cash generated by a project discounting them for the time value of money. If the result of the consolidation of all yearly flow of cash, the Net Present Value (NPV), is positive, the initiative is capable of recovering the initial costs, rewarding the capital invested at an appropriate rate based on the specific risk of the business and generating a surplus.

##### K. Profit & Loss statement

A pro-forma P&L is built to be able to calculate the free cash flow – ideally positive - made available by the commercialization of the vaccine.

- Calculation of Revenues: for each year based on the calculated doses employed (see above) the total revenues are calculated based on the following formula "doses employed x price per dose"
- Calculation of COGS: for each year based on the calculated doses employed (see above) the total costs of the vaccines sold are calculated based on the following formula "doses employed x COGS per dose"
- Calculation of Gross Profit: for each year is calculated as "Revenues – COGS"
- Calculation of Depreciation & Amortization: for each year starting from the year of registration (assumed also as the year of start of depreciation of the costs incurred in the construction of the manufacturing plant), the depreciation amount is calculated as "total cost of manufacturing facility (see below) / depreciation rate (see above)"
- Calculation of SG&A: for each year the total cost incurred for sales force, marketing and administrative expenses is calculated as "total revenues x SG&A rate"
- Calculation of Operating Margin: for each year is calculated as "Gross Profit – Depreciation and Amortization – SG&A"
- Calculation of Income Tax: for each year the income tax due by the company charged with the production and commercialization of the vaccine is calculated as "Operating Margin x Income Tax rate (see above)"
- Calculation of Net Result: for each year the net result is calculated as "Operating Margin – Income Tax"

- Calculation of free Cash Flow: for each year the available cash flow generated by the company is calculated adding back the non-financial items to the Net Result as per the following formula:
  "Net Result + Depreciation and Amortization"

#### L. Clinical development and Manufacturing investments

Costs and cash outflow for all planned clinical trials are also estimated as per report 2
A total cost of 100 million USD is estimated for a greenfield dedicated manufacturing facility.

#### M. NPV Calculation

Once all cash flows are calculated, the Net Present Value (NPV) of the initiative is calculated applying the Weighted Average Cost of Capital (WACC) for the relevant industry (see above) as discount rate as per the following formula:
"$\Sigma_{t=0}^{n}$ Cash Flow$^{t}$ / (1 + WACC)$^{t}$" where t is the number of time periods

---

[i] https://www.who.int/leishmaniasis/burden/endemic-priority-alphabetical/en/
[ii] Pigott et al., Global distribution maps of the leishmaniases, eLife 2014;3:e02851
[iii] Institute for Health Metrics and Evaluation, Global Burden of Disease 2019
[iv] WHO, Report of a meeting of the WHO Expert Committee on the Control of Leishmaniases, Geneva, 22–26 March 2010, p.104
[v] https://www.dndi.org/newsletters/n18/4_1.php
[vi] Zhao-Rong Lun et al., Visceral Leishmaniasis in China: an Endemic Disease under Control, Clin Microbiol Rev. 2015 Oct; 28(4): 987–1004 - In China, three epidemiological types of VL have been described: anthroponotic VL (AVL), mountain-type zoonotic VL (MT-ZVL), and desert-type ZVL (DT-ZVL). AVL is present endemically only in Xinijang where it coexists with MT-VTL. The weight of AVL on the VL total cases in China has been assumed being half of the VL cases in the Xinjinag region as reported in the article.
[vii] Jose A Ruiz Postigo, Leishmaniasis in the World Health Organization Eastern Mediterranean Region, Int J Antimicrob Agents, 2010 Nov;36 Suppl 1:S62-5
[viii] Teleconference with Paul Kaye, August 16, 2019
[ix] Zijlstra et al., Post-kala-azar dermal leishmaniasis in the Indian subcontinent: A threat to the South- East Asia Region Kala-azar Elimination Programme, PLOS Neglected Tropical Diseases, November 16, 2016
[x] Mondel et al., Quantifying the infectiousness of post-kala-azar dermal leishmaniasis towards sandflies, Clin Infect Dis., 2019, Jul 15; 69(2): p.251–258.
[xi] https://www.dndi.org/diseases-projects/leishmaniasis/
[xii] GBD 2017 - Cit.
[xiii] https://apps.who.int/immunization_monitoring/globalsummary/timeseries/tswucoveragedtp3.html
[xiv] Bruni et al., Global estimates of human papillomavirus vaccination coverage by region and income level: a pooled analysis, Lancet Glob Health. 2016 Jul;4(7):e453-63
[xv] https://data.unicef.org/topic/education/primary-education/
[xvi] http://people.stern.nyu.edu/adamodar/New_Home_Page/datafile/wacc.html
